# Cardiac structure and function in schizophrenia: a cardiac MR imaging study

**DOI:** 10.1101/19008649

**Authors:** Emanuele F. Osimo, Stefan P. Brugger, Antonio de Marvao, Toby Pillinger, Thomas Whitehurst, Ben Statton, Marina Quinlan, Alaine Berry, Stuart A. Cook, Declan P. O’Regan, Oliver D. Howes

## Abstract

**Background:** Heart disease is the leading cause of death in schizophrenia.

**Aims:** We investigated cardiac structure and function in patients with schizophrenia using cardiac magnetic resonance imaging (CMR) after excluding medical and metabolic comorbidity.

**Methods:** 80 participants underwent CMR to determine biventricular volumes and function and measures of blood pressure, physical activity, and glycated haemoglobin levels. Patients and controls were matched for age, sex, ethnicity, and body surface area.

**Results:** Patients with schizophrenia had significantly smaller indexed left ventricular (LV) end-diastolic volume, end-systolic volume, stroke volume, right ventricular (RV) end-diastolic volume, end-systolic volume, and stroke volume but unaltered ejection fractions relative to controls. LV concentricity and septal thickness were significantly larger in schizophrenia. The findings were largely unchanged after adjusting for smoking or exercise levels and were independent of medication dose and duration.

**Conclusions:** Patients with schizophrenia show evidence of prognostically-adverse cardiac remodelling compared to matched controls, independent of conventional risk factors.

## Introduction

Schizophrenia is a major mental illness, affecting approximately 0.4-0.7% of the world population(1), equivalent to 30-52.5 million people. Patients with schizophrenia have a 2- to 3-fold greater mortality compared to the general population(2), with a 10 to 25 years shorter life expectancy(3). In addition to the higher suicide risk, mortality due to natural causes has been estimated to be up to 8-times higher than expected(2). Despite reductions in mortality in the general population over the last 30 years (both all-cause and cardiovascular)(4), the mortality gap between patients with schizophrenia and the healthy population is not reducing(5), and may even be increasing(6). Cardiovascular disease (CVD) accounts for a large part of the loss of life in schizophrenia(7) (14.3 life-years lost(8), or 60% of deaths(9), on average) despite being underdiagnosed(10), and its importance has been increasing both in US-based(9) and north-European(4) population samples.

Despite this wealth of epidemiological evidence that associates schizophrenia with cardiovascular disease and premature death, few studies have directly investigated cardiac function in patients with schizophrenia. These have generally shown reductions in left ventricular (LV) ejection fraction and increases in LV mass in schizophrenia(11-13). However, interpretation of these findings is complicated, as the studies did not consistently control for potential confounds, including BMI, gender, and co-morbid metabolic conditions, which are factors that could independently account for the changes that were found. As such, it remains unknown if schizophrenia is associated with cardiac structural or functional alterations, or if potential confounds, such as medical and metabolic comorbidity, explain the differences reported. Furthermore, previous studies have used transthoracic echocardiography (TTE) to assess cardiac function, which provides less accurate and less reproducible volumetric and functional assessment of the heart as compared to cardiac MRI(14).

In view of this, we aimed to determine if cardiac structure and function is altered in patients with schizophrenia using cardiac magnetic resonance (CMR), the gold standard in vivo measure of cardiac structural and functional assessment(15). Using a cohort of patients free of metabolic or medical comorbidities and healthy controls matched for potential confounds including age, sex, ethnicity and body surface area, we tested the hypothesis that patients with schizophrenia show heart changes that could account for the increased cardiovascular mortality independent of established cardiovascular risk factors, such as medical and metabolic comorbidity.

## Methods

### Participants

A total of 40 patients with schizophrenia (mean age 39.8 years; SD 9.6 years) were recruited from South London and the Maudsley NHS Foundation Trust and from Central and North West London NHS Foundation Trust in London, United Kingdom. 40 matched healthy controls (HCs) (mean age 38.9 years; SD 9.4 years) were recruited after matching for age (+/- 3 years), ethnicity, sex, and body surface area (BSA +/- 2, a parameter similar to BMI) through the Hammersmith Hospital Healthy Volunteer Panel, London, United Kingdom and from the UK Digital Heart Project at Imperial College London (volunteers recruited via advertisement as previously described(16)).

Exclusion criteria for all participants were: age <18 or >65 years, pregnancy or breastfeeding, a history of cardiometabolic disease, including diabetes, hypertension, dyslipidaemia, ischaemic heart disease, any vascular disorder, other history of congenital/structural cardiac disease; or history of significant or continuing substance abuse. Inclusion criterion for patients was an ICD-10 diagnosis of schizophrenia. Exclusion criterion for healthy controls was a previous history or first-degree family history of schizophrenia or other psychotic disorder.

Written informed consent was obtained from all volunteers. The authors assert that all procedures contributing to this work comply with the ethical standards of the relevant national and institutional committees on human experimentation and with the Helsinki Declaration of 1975, as revised in 2008. All procedures involving human subjects were approved by the London - Camberwell St Giles Research Ethics Committee.

### Assessment of Participants

Physical assessment, study questionnaires, and cardiac MRI were all performed during the same study visit. All patients were assessed at the time of imaging using the Positive and Negative Syndrome Scale (PANSS) for Schizophrenia(17) by a study psychiatrist. A blood sample for HbA1c was collected from a sub-sample of 38 matched participants (19 patients and 19 controls). Participants were seated in a quiet, temperature-controlled waiting room and given time to relax. A qualified doctor then performed a screening interview, clinical examination, and supervised the MRI scan. The presence of medical co-morbidities was assessed directly by interviewing participants, by examining the list of active prescriptions, and where in doubt via examining general practice records. Brachial blood pressure (BP) measurement was performed following 5 min rest using a validated oscillometric device (Omron M7, Omron Corporation, Kyoto, Japan). The first of 3 measures were discarded, and the second 2 values were averaged to provide the final reading. Physical activity grading was based on the Copenhagen City Heart Study Leisure Time Physical Activity Questionnaire, performed on the same day as the cardiac magnetic resonance imaging. Categories of activity were based on participants’ level of activity over the preceding 12 months, ranging from level 1: almost entirely sedentary, to level 4: >5 hours of exercise per week(18). Plasma HbA1c was analysed using the Tosoh G8 HPLC Analyzer (Tosoh Bioscience, San Francisco). Antipsychotic doses were converted into chlorpromazine equivalents as described by Andreasen and colleagues(19).

### Magnetic Resonance Imaging Protocol

CMR was performed at a single site for all 80 subjects, recruited between September 2015 and December 2018. Due to a hardware upgrade the first 21 patients and matched controls were scanned on a 1.5T Philips Achieva (Best, Netherlands) using a 32-channel cardiac coil and the remaining 19 patients and matched controls on a 3T Siemens Magnetom Prisma (Erlangen, Germany) using a combination of the 18-channel body coil and 12 elements of the 32-channel spine coil. Matched patients and HCs were scanned on the same scanner. All images used a vectorcardiographic technique for ECG synchronisation and were acquired during breath-hold at expiration. A standard clinical protocol for assessing biventricular function and volumes was followed according to published international guidelines(20). Subjects whose images were degraded by respiration or ECG synchronisation artefacts to the extent where cardiac contours could not be clearly identified were excluded from the analysis. Images were stored on an open-source database(21). Volumetric analysis of the cine images was performed using CMRtools (Cardiovascular Imaging Solutions, London, UK) by experienced operators blind to the diagnosis. The epicardial and endocardial borders were manually contoured at end-diastole and end-systole in each of the short-axis slices. Semi-automated, signal intensity-based thresholding was then used to include the papillary muscles in the left ventricular mass and excluded from volumetric measurements. To increase the accuracy of the analysis, the valve positions at end-diastole and systole were manually identified on the three long-axis images, allowing the valve planes to be tracked through the cardiac cycle (Figure 1). Volumes and mass were indexed to body surface area calculated using the Mosteller formula (Height [cm] × Weight [kg]/3600)^½^. Indexed volumetric data were left ventricular (LV) mass (LVMi), LV and right ventricular (RV) end-diastolic volumes (LVEDVi and RVEDVi), LV and RV end-systolic volumes (LVESVi and RVESVi), and LV and RV stroke volumes (LVSVi and RVSVi). The end-diastolic volume (EDV) and end-systolic volume (ESV) were calculated using the Simpson’s method(22). Stroke volume (SV) was derived from EDV-ESV and ejection fraction by SV/EDV x 100. Left ventricular mass was the total epicardial volume minus the total endocardial volume multiplied by the specific density of the myocardium (1.05 g/ml). Maximal end-diastolic wall thickness was measured through the basal antero-septal wall using a short-axis CMR image (see figure 1C). LV concentricity was calculated as LV mass to LV EDV ratio. Pulse-wave velocity (PWV) was calculated using the method described in (23).

**Figure 1:**
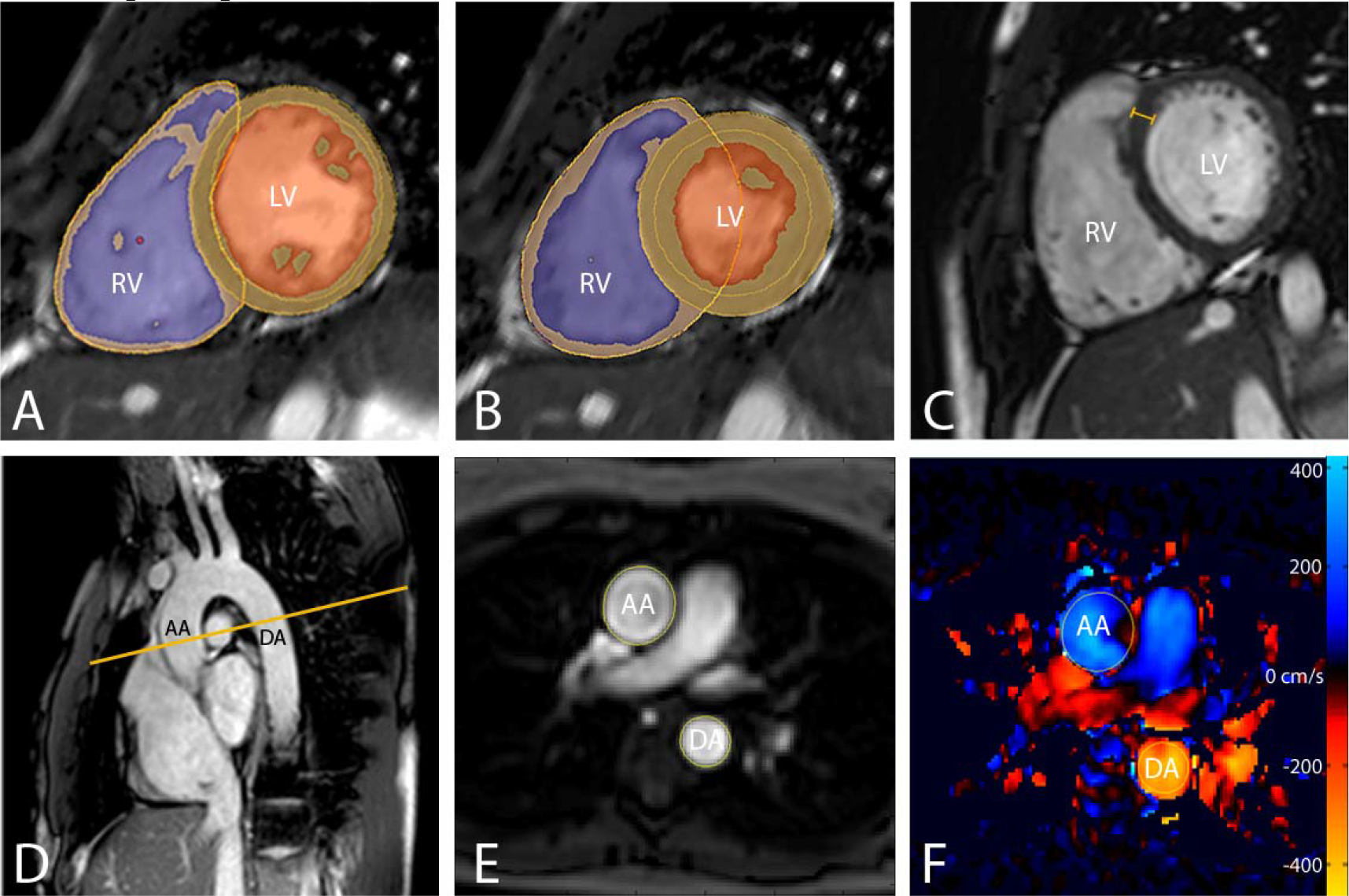
MR images demonstrating the assessment of biventricular volumes and function in an adult participant. (A & B) <u>Ventricular Function and Mass</u> Endo- and Epicardial contours of the left and right ventricle in diastole (A) and systole (B). Myocardium segmentation shown by orange lines, left ventricular cavity shown in orange and right ventricular cavity shown in purple. For ventricular function assessment, the endocardial and epicardial contours of the left and right ventricle were delineated in diastole (A) and systole (B) on a stack of short axis slices covering the entire ventricles. (C) <u>Septal Thickness measurement shown by the orange calliper</u> (D, E & F) <u>Pulse Wave Velocity</u> Transverse CMR slice through the aortic arch (D) displaying the magnitude (E) and velocity encoded (F) images through the aortic arch. Yellow contours outline the ascending aorta (AA) and descending aorta (DA). Blood flow through the aorta is encoded with a signal intensity relative to its velocity (cm/s), shown here on a blue/red/yellow colour scale (F).

### Statistical analysis

Differences among patients and controls were tested using χ^2^ tests for categorical variables, Wilcoxon rank sum tests for non-normally distributed values, and analysis of variance (anova) for normally distributed measures. Effect sizes were calculated using Cohen’s *d* measure.

As there were significant group differences in smoking and physical activity levels, sensitivity analyses were conducted using generalised linear models with smoking, physical activity levels, and scanner (1.5T or 3T) as covariates, the cardiac measures as the dependent variables and group as the independent variable.

In order to explore the possibility of a dose-response relationship between medication and cardiac phenotypes, linear regression was used to correlate cardiac measures to the natural logarithm of total chlorpromazine equivalents (to normalise the data distribution). The overall proportion of variance explained by linear models was calculated using R^2^ measures.

In all tests, a p value <0.05 (two-tailed) was taken as significant, and p values were adjusted for multiple testing using the Benjamini & Hochberg method. Statistical analyses and graph plotting were performed in R (R Foundation for Statistical Computing, 2019, Vienna, Austria).

## Results

A total of 80 subjects were recruited for this study. One healthy control had to be excluded due to poor image quality (Table 1). Subject characteristics are described in Table 1. At the time of assessment, all patients were treated with antipsychotics (clozapine; N=28, olanzapine; N=10 and risperidone N=2). There were no significant group differences for age, sex, ethnicity, BSA, HbA1c, systolic and diastolic blood pressure. However, smoking levels and weekly physical activity levels were significantly different between groups.

**Table 1:** Sample Characteristics. IQR: inter-quartile range; BSA: body surface area (calculated using Mosteller formula); BMI: body mass index; BP: blood pressure; SD: standard deviation; df: degrees of freedom; HbA1c: glycated haemoglobin

| Characteristic | Schizophrenia | Healthy Controls | Test statistic and p value |
| --- | --- | --- | --- |
| **Sample size** | 40 | 39 |  |
| **Male sex, N (%)** | 31 (77.5%) | 32 (82.0%) | $\chi^2 = 0.2$; d.f. = 1; p=0.61 |
| **Caucasian ethnicity, N (%)** | 19 (47.5%) | 21 (53.9%) | $\chi^2 = 0.32$; d.f. = 1; p=0.57 |
| **Age, years mean (SD)** | 39.80 (9.55) | 38.92 (9.38) | F= 0.17, df=1, p=0.68 |
| **Number of cigarettes smoked per day, median (IQR; min; max)** | **1 (4.25; 0; 35)** | **0 (0; 0; 10)** | **Wilcoxon rank sum test**<br>**p<0.001** |
| **Activity score, median (IQR)** | **2 (0)** | **3 (1.5)** | **Wilcoxon rank sum test**<br>**p<0.001** |
| **BSA (m<sup>2</sup>), mean (SD)** | 2.05 (0.27) | 1.95 (0.22) | F=3.45, df=1, p=0.07 |
| **Systolic BP (mmHg), mean (SD)** | 124.64 (11.11) | 122.05 (12.45) | F= 0.90, df=1, p=0.35 |
| **Diastolic BP (mmHg), mean (SD)** | 81.22 (9.50) | 77.78 (9.58) | F= 2.44, df=1, p=0.12 |
| **HbA1c (mmol/mol), mean (SD)\*** | 36.28 (6.02) | 33.90 (7.92) | F=1.04, df=1, p= 0.31 |
| **Chlorpromazine Equivalent Dose (mg/day), Median (IQR)** | 377 (290) | 0 (NA) | NA |
| **Duration of treatment (years), Median (IQR)** | 12 (13) | NA | NA |
| **Total PANSS score, Median (IQR)** | 62 (35.5) | NA | NA |

\* A blood sample for HbA1c was collected from a sub-sample of 38 matched participants (19 patients and 19 controls).

### Cardiac Measurements of Ventricular Structure and Function in Patients with Schizophrenia and Matched Healthy Controls

Supplementary Table 1 and Figures 2-4 describe CMR-derived measures of cardiac volume and function according to diagnostic status. After adjusting results for multiple testing, patients with schizophrenia had significantly smaller indexed LV end-diastolic volume (*d*=-0.82, F=-11.90, p=0.001), LV end-systolic volume (*d*=-0.58, F=-4.65, p=0.02), LV stroke volume (*d*=-0.85, F=-7.25, p=0.001) (Figure 2), RV end-diastolic volume (*d*=-0.79, F=-15.32, p=0.002), RV end-systolic volume (*d*=-0.58, F=-7.36, p=0.02), and RV stroke volume (*d*=-0.87, F=-8.09, p=0.001) with large effect sizes (Figure 3). LV concentricity (*d*=0.73, F=0.12, p=0.003) and septal thickness (*d*=1.13, F=1.71, p<0.001) were significantly larger in people with a diagnosis of schizophrenia compared to matched healthy controls with large to very large effect sizes (Figure 4). There were no significant differences between groups in LV mass, biventricular ejection fractions, or in pulse-wave velocity (Supplementary Table 1 and Supplementary Figure 1).

**Figure 2:**
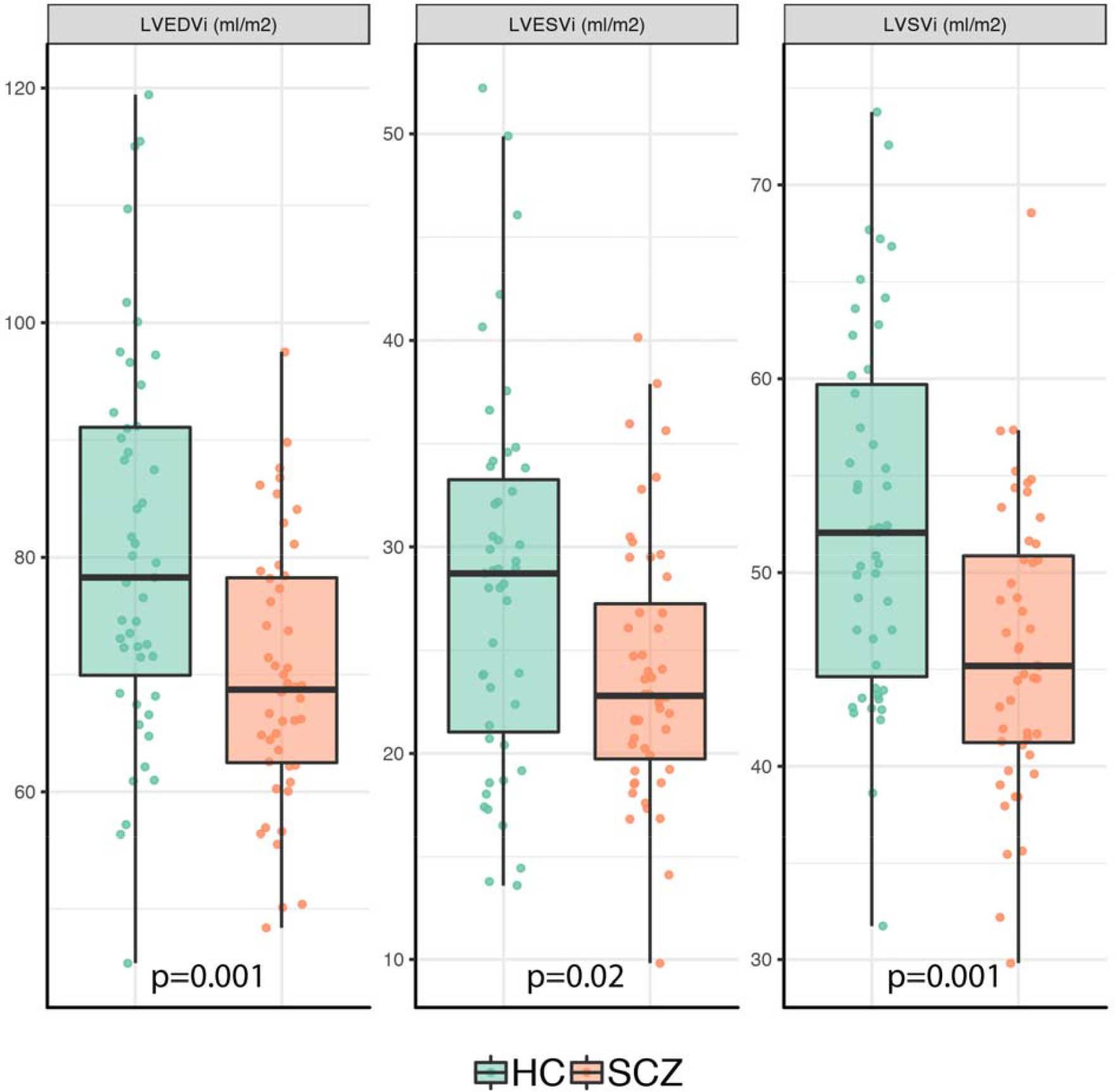
Left Ventricular Cardiac Measurements in Patients with Schizophrenia and Healthy Controls. Graphs show individual values and box and whisker plots (the solid horizontal line is the median, the lower and upper hinges correspond to the first and third quartiles (the 25th and 75th percentiles), and the whiskers extends from the hinge to the largest/smallest value no further than 1.5*inter-quartile range from the hinge). P values are adjusted for multiple testing. Patients with schizophrenia had significantly smaller indexed LV end-diastolic volume, indexed LV end-systolic volume, indexed LV stroke volume

**Figure 3:**
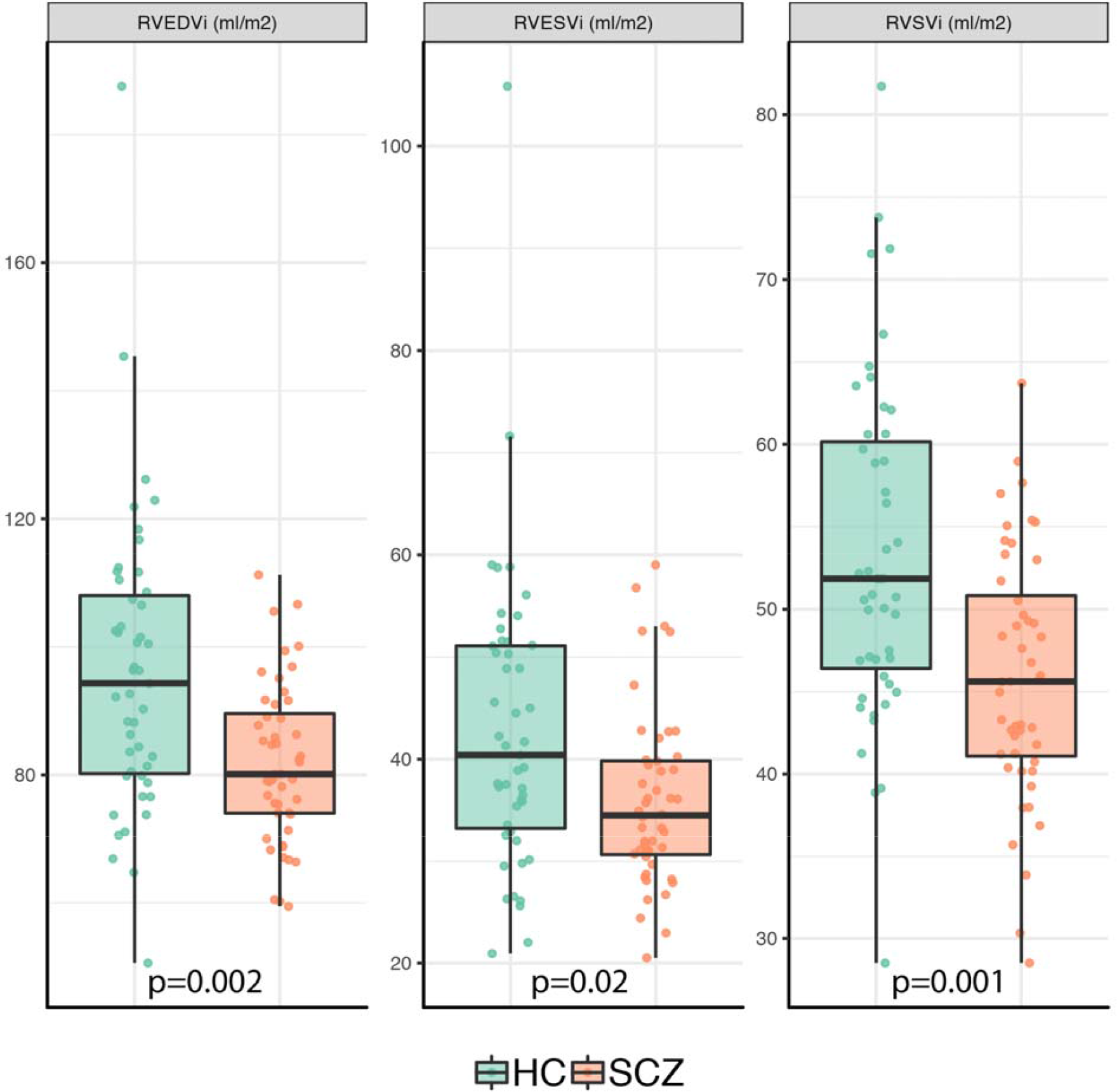
Right Ventricular Cardiac Measurements in Patients with Schizophrenia and Healthy Controls. Graphs show individual values and box and whisker plots (the solid horizontal line is the median, the lower and upper hinges correspond to the first and third quartiles (the 25th and 75th percentiles), and the whiskers extends from the hinge to the largest/smallest value no further than 1.5*inter-quartile range from the hinge). P values are adjusted for multiple testing. Patients with schizophrenia had significantly smaller indexed RV end-diastolic volume, indexed RV end-systolic volume, and indexed RV stroke volume.

**Figure 4:**
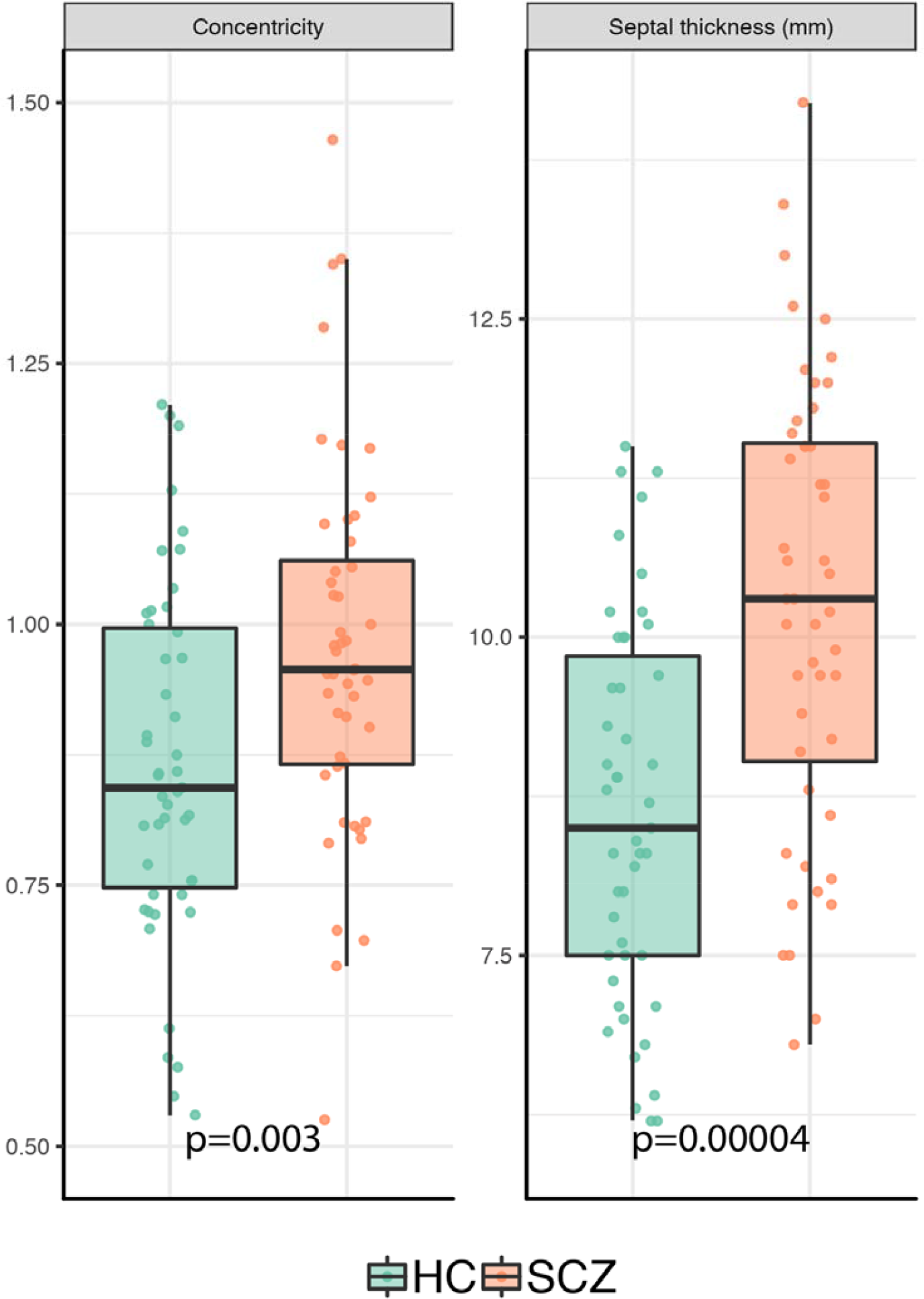
Other Cardiac Measurements in Patients with Schizophrenia and Healthy Controls. Graphs show individual values and box and whisker plots (the solid horizontal line is the median, the lower and upper hinges correspond to the first and third quartiles (the 25th and 75th percentiles), and the whiskers extends from the hinge to the largest/smallest value no further than 1.5*inter-quartile range from the hinge). P values are adjusted for multiple testing. LV concentricity and septal thickness were significantly larger in people with a diagnosis of schizophrenia compared to matched healthy controls.

#### Sensitivity analyses

In sensitivity analyses, we explored whether significant differences between patients and controls in terms of cigarettes smoked per day and number of hours of weekly exercise contributed to the differences in cardiac measurements we observed. Supplementary Table 1 shows that, after adjusting the model for the number of cigarettes smoked per day, patients with schizophrenia still exhibited significantly smaller LVEDVi (p=0.004), LVESVi (p=0.04), LVSVi (p=0.003), RVEDVi (p=0.006), RVESVi (p=0.04), and RVSVi (p=0.003), while LV concentricity (p=0.02) and septal thickness (p<0.001) were both still significantly larger. After co-varying for physical activity levels, patients with schizophrenia exhibited significantly smaller LVEDVi (p=0.04), LVSVi (p=0.04), RVEDVi (p=0.04), RVSVi (p=0.04), while LV concentricity (p=0.04) and septal thickness (p<0.001) were both significantly larger. Adjusting for scanner (1.5 vs 3T) did not affect the significance of any of the results (Supplementary Table 2).

#### Associations Between Cardiac Measures, Total Medication Dose and Duration of Treatment

In linear regression analyses, no cardiac measure significantly correlated with total current chlorpromazine dose equivalents (Supplementary Table 3), nor with total chlorpromazine equivalents/years (Supplementary Table 4).

## Discussion

In this study we found that patients with schizophrenia have clinically significant reductions in indexed end-diastolic, end-systolic and stroke volumes in both ventricles (with large effect sizes), while showing an increase in concentricity and septal thickness (with large to very large effect sizes), when compared to healthy controls of a similar age, sex, ethnicity, body surface area and blood pressure. These findings were unchanged in sensitivity analyses adjusting for smoking levels or exercise levels, other than biventricular end-systolic volumes, which were no longer significantly different. In other words, for the first time to our knowledge, this study shows evidence consistent with concentric remodelling of the heart in schizophrenia. Cardiac remodelling is the response of the heart myocytes and vasculature to noxious stimuli that leads to structural changes, a process that can become pathogenic(24). Specifically, concentric remodelling is defined as that happening when the EF is preserved, volumes are normal or reduced, but the left ventricular mass to volume ratio is increased(24). Furthermore, the changes found in our sample of individuals with schizophrenia were independent of conventional cardiometabolic risk factors such as differences in age, sex, ethnicity, medical comorbidity, BSA, HbA1c levels or blood pressure; something that previous studies(11-13) could not consistently exclude, and could therefore account for the difference in results.

### Strengths and Limitations

A strength of this study is the use of CMR, which is the gold standard for LV function and mass quantification(15). A key advantage of CMR over echocardiography is the ventricles can be imaged in their entirety, and no geometrical assumptions need to be made in order to derive global whole organ data(14). Another strength of our study is that, for the first time to our knowledge, participants with a pre-existing diagnosis of any medical condition, including ischaemic heart disease, hypertension, diabetes, or dyslipidaemia were excluded, and we matched patients and controls for BSA, age, sex and ethnicity, all potential confounders in analyses of cardiac function and structure(15, 25-27). However, whilst we excluded diagnosed cardiometabolic disorders, we cannot exclude the possibility of sub-clinical changes, although we found no significant difference between cases and controls in systolic or diastolic blood pressure, and no significant difference in HbA1c in a sub-sample of our cohort, suggesting sub-clinical differences are unlikely to be large. A limitation of this study is that it involved chronic patients (median duration of treatment of 12 years), who were all taking antipsychotic medications. Thus, a future study in antipsychotic naïve patients would be useful to determine if the cardiac differences we detect are associated with treatment, or chronicity. Another consideration is that subjects were scanned on two different scanners. However, findings remained significant when scanner was included in the analysis. Moreover, recent evidence shows that 1.5T or 3T scanners yields similar results for measuring cardiac structure and function(28), suggesting the change in equipment is unlikely to have had a major effects on our results.

### Implications of our findings

Concentric cardiac remodelling has been shown as the strongest predictor of future cardiovascular disease events such as myocardial infarction, coronary insufficiency, heart failure, and stroke in healthy adults(29), and demonstrated the lowest CVD-free survival over time and steepest slope curves among other measures such as LV mass and relative wall thickness, with a hazard ratio of 1.40(29). Increased LV septal thickness also confers higher cardiovascular morbidity and mortality(30). The cardiac changes we report could therefore account for part of the increased cardiovascular morbidity and mortality seen in schizophrenia, independent of the increased cardiovascular risk conferred to patients with schizophrenia by conventional risk factors due to secondary effects of the illness (e.g. a sedentary lifestyle(31), higher rates of smoking, the development of metabolic syndrome linked to the metabolic consequences of some antipsychotic treatments(32, 33), and altered glucose(34) and lipid homeostasis(35)).

Concentric cardiac remodelling in schizophrenia, independent of conventional cardiometabolic risk factors, could be caused by either direct effects of antipsychotic treatment, or linked to the pathophysiology underlying the illness, such as inflammation(36). Schizophrenia has been associated with elevations in peripheral pro-inflammatory cytokines, including in interleukin-6 (IL-6) and tumour necrosis factor levels in antipsychotic-naïve individuals(37). IL-6 and related cytokines play a crucial role in the regulation of cardiac myocyte hypertrophy and apoptosis(38), and initial evidence supports an early diffuse fibro-inflammatory myocardial process in patients with schizophrenia(39).

With regards to antipsychotic treatment, there is direct animal evidence that dopamine receptor occupancy contributes to regulating cardiac function, and that antipsychotics impair this by blocking D2/3 receptors(40), thus potentially contributing to cardiac fibrosis(41). Antipsychotics are also associated with the development of inflammatory lesions and lymphocytic infiltrates within the myocardium(42). Therefore, antipsychotic treatment could account for some of the cardiac changes we demonstrate. However, in our sample we did not find a linear relationship between cardiac changes and current or lifetime antipsychotic dose. The absence of a relationship between them reduces the probability that a causal relationship exists(43), even if this cannot be ruled out. Future studies will need to address whether cardiac changes in schizophrenia are due to illness-specific effects, or to antipsychotic treatment, or both.

## Conclusions and Future Directions

We observed biventricular volume reductions and evidence of concentric cardiac remodelling in patients with schizophrenia as compared to matched healthy controls in the absence of concurrent established metabolic or medical risk factors. These are prognostically adverse changes, and, in the absence of concurrent conventional metabolic or medical risk factors, could explain part of the additional cardiovascular morbidity and mortality seen in schizophrenia. Further work involving untreated subjects is required to disentangle the contribution of antipsychotic medication on cardiac changes in schizophrenia.

## Data Availability

The dataset for this paper is available on Code Ocean together with the R analysis codes, which can be reproduced on the go (https://doi.org/10.24433/CO.9265392.v1).

## Acknowledgments, contributions and funding

EFO and SB contributed to the design of the study, recruited and scanned participants, performed data analyses, and drafted the manuscript. AdM, BS, MQ, and AB contributed to study design, cardiac image analysis, and revised the manuscript. TW and TP contributed to participant recruitment and scanning and revised the manuscript. SAC revised the manuscript. DPOR supervised data analyses and revised the manuscript. ODH conceived and designed the study, supervised data analyses, and revised the manuscript. All authors have approved the final manuscript. We would like to thank Alain DeCesare and Pawel F. Tokarczuk for their help in analysing PWV data.

This study was funded by grants MC-A656-5QD30 from the Medical Research Council-UK, and the NIHR Biomedical Research Centre South London and Maudsley Foundation NHS Trust to Professor Howes.

